# Evaluating Online Educational Resources for Osteogenesis Imperfecta: Insights from Google, Bing, and ChatGPT3.5

**DOI:** 10.1101/2025.02.09.25321967

**Authors:** Victoria-Jessica C. Partin, Andrea M. Hooberman-Pineiro, April A. Pace

## Abstract

**Background:** Osteogenesis Imperfecta is a rare connective tissue disorder resulting in a diverse range of musculoskeletal deformities. Patient education is an important aspect of medical care, and the internet is a popular place in which patients seek information about their medical conditions. This is a cross-sectional study aimed to evaluate the quality of online patient educational websites about Osteogenesis Imperfecta.

**Methods:** The authors searched for patient education websites using Google and Bing. Twenty patient education websites about Osteogenesis Imperfecta were collected from each. Websites meeting inclusion criteria were evaluated using the DISCERN tool, JAMA Benchmark Criteria evaluation, and a Flesch-Kincaid Readability test. T-tests were used to compare search engine results. Websites were also collected from ChatGPT3.5, however, due to hallucinations and exclusion criteria, the sample size was too small to compare to the search engines.

**Results:** No significant differences were found between search engines, and the quality of the websites was not high. Average overall ratings for DISCERN were mediocre, and for the JAMA criteria they were low. The average readability scores required 8-9 grade level reading comprehension, although there was variability within each search engine.

**Conclusion:** Neither search engine provided significantly better sources than the other, the overall quality of the websites was not high, and the low readability scores could make these difficult for patients with lower health literacy to understand

## Background

Osteogenesis imperfecta (OI) is a rare autosomal recessive connective tissue disorder that results in a variety of systemic musculoskeletal deformities. The severity of OI ranges from normal appearing individuals who experience joint hypermobility or a series of fractures in their life, to people who live with more severe disabilities such as hearing loss or cardiopulmonary complications, and some may even rely on wheelchair assistance. The prevalence of OI is equal amongst race, ethnicity, and sex (1).

Patient education is an important aspect of medical care, and it is crucial that physicians are well versed in what websites patients may have access to online. Often, when receiving a diagnosis, patients and parents will quickly resort to researching information on the internet. While the internet provides an extensive amount of resources, not every website may be high quality or reliable. Additionally, health literacy is another key factor to patient care. Only 12% of Americans are proficient in health literacy and an overwhelming majority of people who are below proficiency are from vulnerable and underserved populations (2). This poses a concern that patients who have OI may not understand the disease or treatment options as well, impacting their quality of care. This project aimed to review the quality of educational material presented online regarding OI, based upon multiple criteria, including reliability and readability. This was accomplished by applying the DISCERN Instrument, JAMA Benchmark Criteria, and Flesch-Kincaid Readability test to a list of websites provided on Google, Bing, and ChatGPT 3.5. The use of multiple instruments and criteria are important in the evaluation of patient education, considering the range in health literacy within the patient population

The DISCERN Instrument is a questionnaire that provides individuals a credible and consistent approach to assessing the quality of written information on treatment options for a health condition (3). This tool allows individuals to think critically about the treatment information presented on a website. The JAMA Benchmark Criteria uses four core standards to evaluate readability, and the Flesch-Kincaid Readability test assesses reading ease and approximate grade level required for understanding of the websites (4,5). These two tools are important because not all patients have the same educational background, and it is critical to present information in a way that is easily digestible.

In addition, the use of artificial intelligence (AI) large language models is trending; a notable popular one is ChatGPT3.5. While the exact usage of AI in patient education is unclear, it is not unreasonable to assume that patients may be using these platforms to look for answers due to AI’s prevalence and accessibility (6). While ChatGPT3.5 can seem to know all, it is also known to have faults. It can come up with responses by making false assumptions, and while they may seem dependable, answers can be incorrect and made up; these invented answers are often termed hallucinations (7). Hallucinations can pose a risk to the quality of a patient’s health because not all the information provided on the platform may be accurate or up to date. ChatGPT3.5 is the version available for free, and this model does not always have the most up to date information to draw from (8). The more updated version is available with a paid subscription. This is a limitation to the platform as medical knowledge continues to evolve quickly. Therefore, it is important that these AI informational sources are also being evaluated when it comes to patient education and healthcare. Because of this, it was pertinent to include ChatGPT3.5 in this evaluation of patient education on osteogenesis imperfecta.

This study aimed to evaluate the quality and readability of patient education websites on Osteogenesis imperfecta through Google and Bing search engines, and a large language model (ChatGPT3.5). Sources were evaluated from both Google and Bing and were compared. Websites from ChatGPT were evaluated but were not included in the comparison due to the small sample size. Through analysis of the websites provided gaps were identified and recommendations were developed regarding patient education on websites.

## Methods

The term “Osteogenesis Imperfecta” was searched in Google and Bing. This was done through incognito mode to avoid proximity bias in the search engines (9). The first twenty websites from each search engines were selected and reviewed against the exclusion criteria: website requires a subscription, there was a duplicate website within the list, website was in a different language, or ones that did not have patient education information to review. Overall, two websites were excluded from Google and six were excluded from Bing.

The question “can you provide me with 20 patient education references for osteogenesis imperfecta” was used when searching for websites in ChatGPT3.5. It was not possible to use the same methods as was done for Google and Bing because ChatGPT3.5 simply provided a description of OI rather than websites. The list of websites provided was reviewed against the same exclusion criteria in addition to excluding any hallucination websites (10). Overall, sixteen websites were excluded from ChatGPT3.5.

### DISCERN

The Discern analysis tool is a questionnaire that consists of a series of sixteen questions that provides users with a reliable way to evaluate information regarding treatment choices presented within a website (3). Eighteen websites from Google, fourteen websites from Bing, and four websites from ChatGPT3.5 were analyzed against the DISCERN tool by two independent reviewers.

### JAMA

The JAMA benchmark criteria uses four core standards (authorship, attribution, disclosure, and currency) to evaluate the validity of websites. A total of eight questions are asked in a binary fashion and websites are scored on a scale of zero to four (4). Eighteen websites from Google, fourteen websites from Bing, and four websites from ChatGPT were analyzed against the JAMA criteria by two reviewers.

### FLESCH-KINCAID

The online calculator Flesch-Kincaid was used to determine the Grade Level (GL) and Reading Ease (RE) of the websites used in this study. GL was analyzed by the average word complexity and sentence length of each website. The RE score is ranked on a scale from 1 to 100, with 100 being the highest score. A total of eighteen websites from Google, fourteen websites from Bing, and four websites from ChatGPT3.5 were analyzed against the Flesch-Kincaid criteria by two reviewers (4,5).

### Data Analysis

A two sample T test assuming equal variance was used to analyze the DISCERN, JAMA, and Flesch-Kincaid scores between Google and Bing. Data analysis was not conducted on ChatGPT 3.5 due to its small sample size. Additionally, a weighted kappa was used to determine any variability in the DISCERN scores between the reviewers. Below is a summary detailing the kappa score in relation to strength of agreement amongst reviewers.

## Results

### DISCERN

In the Google and Bing search results, some websites scored well, and others poorly based on the DISCERN tool. For Google, out of 18 included websites, two received the highest average rating of 5, and two scored the lowest at 1. For Bing, among 14 included websites, one achieved the highest score of 5, and four received the lowest score of 1. The average overall ratings were 2.61 for Google and 2.50 for Bing (on a scale of 1-5). When these mean DISCERN ratings were compared, the difference between the search engines was not statistically significant (p=0.8131) (see Table 2).

To measure possible variability between how the reviewers scored the websites, a kappa was performed by analyzing DISCERN’s final question (question 16) regarding overall rating of the website, and between all the DISCERN questions (Table 3). There was moderate strength of agreement for Google, and a good strength of agreement for the Bing websites (Table 1). For both Google and Bing, the combined agreement for all questions 1-16 had moderate strength of agreement.

**Table 1.**
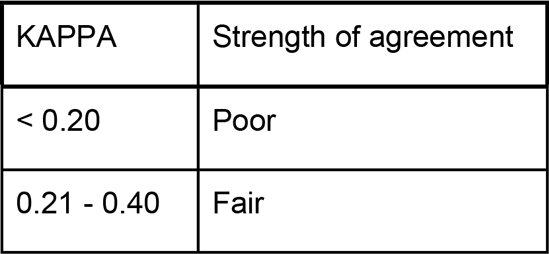
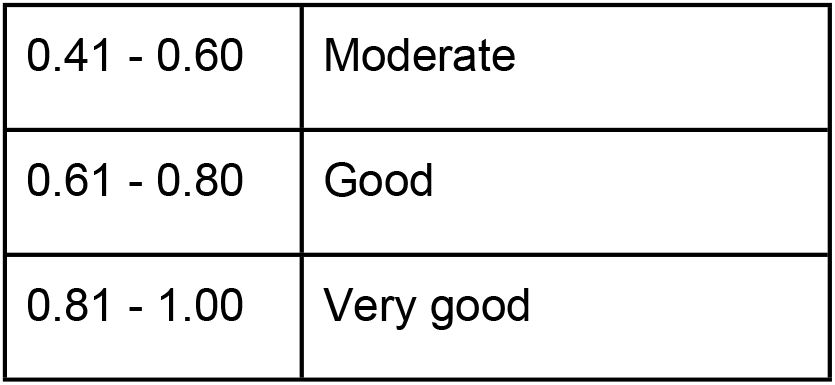

**Table 2.** Equal variance two-tailed t-test values between Google and Bing.

|  | **DISCERN**<br>**(overall**<br>**question 16)** | **DISCERN**<br>**(questions**<br>**1-15)** | **JAMA** | **Flesch-Kincaid**<br>**Reading Ease** | **Flesch-Kincaid**<br>**Grade Level** |
| --- | --- | --- | --- | --- | --- |
| **P value** | **0.813** | **0.989** | **0.617** | **0.482** | **0.345** |

**Table 3.**
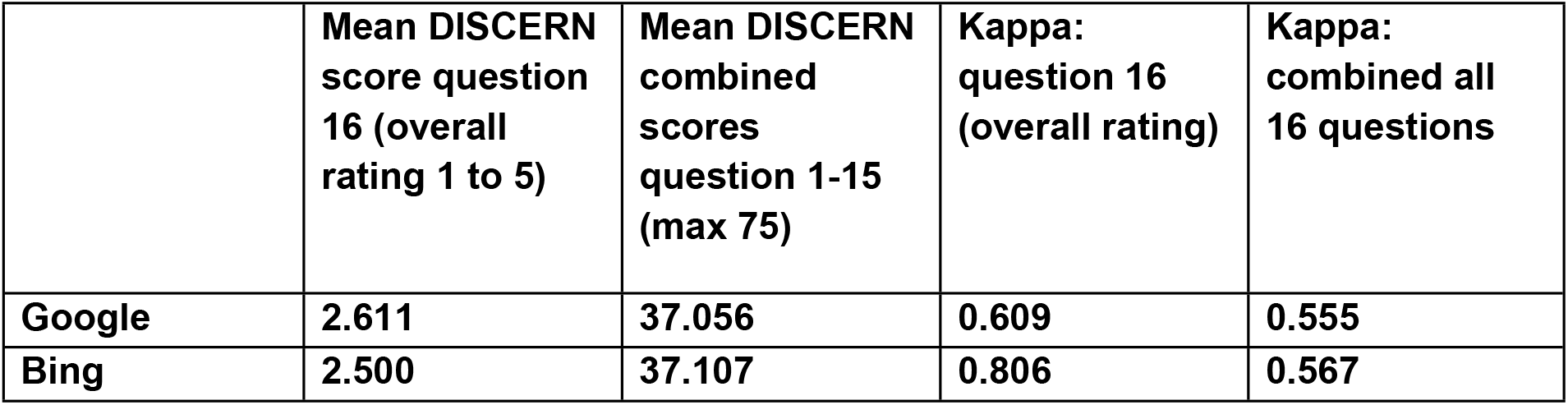
DISCERN average scores and Kappa comparison between reviewers.

### JAMA

For the JAMA Benchmark Criteria scores, rated on a scale from one to four, the means for both Google and Bing did not differ significantly (p=0.6165, Table 4). While there was no significant difference overall, the two highest scoring websites, each achieving the top rating of 4, were found on the Bing search engine. However, even with these scores, the average Bing rating was only 1.21, and the average Google rating was 1.00.

**Table 4.** JAMA Benchmark criteria average scores.

|  | **JAMA mean score** |
| --- | --- |
| **Google** | **1.000** |
| **Bing** | **1.214** |

### FLESCH-KINCAID

Overall, the Flesch Kincaid Reading Ease (RE) and Grade Level (GL) scores (Table 5) were not significantly different between Google and Bing (p = 0.478696 and 0.344165 respectively). While the data was insignificant, it was noted that one website from Bing had the lowest overall Reading Ease score of 17.4, equivalent to a grade level of 12.4. In comparison, Google’s lowest Reading ease score was 28, equivalent to a grade level of 10.2. Additionally, Google had four websites that scored in the 60s with the highest score being 65.9, equivalent to a grade level of 5.2, while Bing only had one website score in the sixties at 62.5, equivalent to a grade level of 6.3.

**Table 5.** Flesch-Kincaid Average Scores.

|  | **Mean Flesch-Kincaid Reading Ease** | **Mean Flesch-Kincaid Grade Level** |
| --- | --- | --- |
| **Google** | **45.839** | **8.556** |
| **Bing** | **42.779** | **9.136** |

### CHATGPT3.5

The data collected from ChatGPT3.5 resulted in an average DISCERN score of 2.5, average JAMA score of 1.25, an average reading ease of 53.625, and an average grade level of 7.375 (Table 6). However, after exclusion criteria, there were only four websites that met the inclusion criteria and were not hallucinations, and therefore the sample size was too small to conduct a more thorough data analysis.

**Table 6.** ChatGPT data means.

|  | **DISCERN**<br>**(question 16)** | **JAMA** | **Flesch-Kincaid**<br>**Reading Ease** | **Flesch-Kincaid**<br>**Grade Level** |
| --- | --- | --- | --- | --- |
| **ChatGPT** | **2.5** | **1.25** | **53.625** | **7.375** |

## Discussion

Osteogenesis Imperfecta (OI), also known as brittle bone disease, is an autosomal recessive disorder that impacts the skeletal muscle system, leading to low bone mass and increased bone fragility. OI is commonly diagnosed in infancy or childhood and is a lifelong disorder that often results in pain, immobility, and overall skeletal deformities (11,12).

Connective tissue integrity is largely impacted by OI, manifesting in a range of symptoms such as blue sclera, hearing loss, and joint hypermobility. Because there are over nineteen types of OI, every patient is impacted differently and requires different forms of treatment and care (1).

Often when a child is diagnosed with a disease such as OI, parents will look for information online to best understand the diagnosis, treatments, and how those will impact overall quality of life for their child. Therefore, patient education is a crucial aspect of healthcare for patients and their parents to feel empowered in their decision-making regarding treatment options. Search engines such as Google and Bing are tools commonly used for patient education. With the introduction of ChatGPT3.5, patients now have another educational tool at the tip of their fingers. As physicians, it is important to understand the quality of health information that is presented online and through artificial intelligence (AI).

After evaluating each website using the DISCERN tool, neither search engine provided better articles than the other, and there was variation within each search engine’s results. This may provide difficulties for patients when they are trying to sift through websites on their own, due to the variability in quality of sources. However, it is worth noting that websites that did not discuss treatments scored lower on this scale due to reviewers not being able to rate treatment related questions in the DISCERN tool. This would lower an article’s overall score regardless of the quality of information that was presented. A kappa was performed due to the possibility of reviewer biases when rating the sources, a potential limitation of the study. However, variability in kappa scores could also reflect the strength of the sources, as having lower agreement in answering the DISCERN questions could indicate a lack of clarity within the article.

Using the JAMA benchmark criteria, which focuses on reliability and transparency, there was no significant difference between the two websites, with both Google and Bing evaluating poorly when using this tool This suggests indicates that the information may have issues regarding reliability.

Reading Ease is scored on a scale of 1 to 100, with 100 being the highest readability score, therefore the higher the score the easier it is for patients to read a website. The score is determined by reviewing the sentence length and word length of an article. A score of 60-70 is equivalent to grades 8-9 and therefore should be easily read and understood by most individuals aged 13-15 years old. This data shows that despite the insignificant results, Google had a larger number of websites in the easier readability range as compared to Bing. It is important to distinguish which search engine has a higher density of easily readable websites as interpreting health language and information are additional steps that patients are required to take. Removing any literary roadblocks is a crucial step of patient care.

Health literacy is an important component of maintaining a healthy lifestyle as well as enabling individuals to make informed decisions about health care and medical treatment.

Currently, 12% of Americans are proficient in health literacy, while 14% are below basic understanding. The overwhelming majority of patients who are not proficient are more likely to come from a minority group. Data in 2024 identifies that 24% of African American adults, 41% of Hispanic adults, and 25% of American Indian and Native Alaskan adults have a below understanding of basic health literacy. Additionally, 56% of people aged 75 and older are below health literacy (2).

There are some general observations made from the review of patient education websites available for Osteogenesis Imperfecta. One of the major factors that lowered many scores within the DISCERN tool was a lack of in-depth information. Many websites received lower scores due to a lack of information regarding treatments. There were many websites regarding general benefits and risks, but not in-depth discussions regarding benefits and risks of each specific treatment. This is important as, especially with OI, there are many different treatment options and different healthcare specialists involved. The DISCERN question “Does it describe what would happen if no treatment is used?” consistently had low scores across websites. This is an important question for many patients, especially considering barriers to healthcare that exist, including but not limited to costs, transportation, and time. Therefore, more information regarding benefits, risks, and outcomes of no treatment would greatly improve patient education. These factors are not limited to OI and should be considered across other disorders and diagnoses as well if not already addressed.

The second observation is that many of the websites did not have a statement referring to the aims of the websites. Many had low scores when it came to the DISCERN question regarding clear aims. This is important to help guide readers, and the absence of these clear aims likely means additional time spent working through which websites are useful. A potential solution would be to have bullets listed in the beginning to help inform readers when first opening the webpage about what information will be available on the website.

Thirdly, the scores for both Google and Bing when using the JAMA criteria were low. The JAMA criteria assess an article’s reliability, and most websites were unable to meet these criteria including questions about authorship transparency, references listed, website/organization transparency, and currency of the information. Many foundation and hospital websites did not have sources listed, and there was a lack of links to additional information, including to information outside of an organization’s own website. These areas need overall improvement for patients to have reliable websites available. Until these changes occur, it is important for physicians to help educate their patients on how to look for reliable information online, so that patients are aware of the possible inaccuracies or biased information they may come across.

### ChatGPT3.5

AI large language models are growing in popularity. Therefore, it was important to include a common one (ChatGPT 3.5) in this study. When asked to generate patient education references, ChatGPT3.5 was not able to produce many. The prompt used for ChatGPT3.5 asked for 20 references, however only 4 results included links that lead to actual content, the rest were hallucinations. The home websites were often real, however the articles on those websites were not. For example, most of the links led to “page not found” errors. In addition, of those 4 websites with working links, 3 were also found on Google and/or Bing. Due to the limited sample size, a thorough data analysis on these websites was not conducted.

It is acknowledged that most people will directly input their questions in the search bar on ChatGPT, however, to evaluate patient education found through ChatGPT in a similar manner as Google and Bing, ChatGPT was asked to provide OI references. The amount of hallucinations present in the ChatGPT response is also an indicator that there are still major limitations to ChatGPT, and this continues to raise the question of reliability. Generic large language models, like ChatGPT, may not be the best method at this time for finding patient education websites.

### Recommendations

Based on findings and observations several recommendations can be made regarding how developers create online content for patient education.

- The first is to create in-depth content regarding each specific treatment, including a section regarding information on outcomes with no treatment.
- The second is to have a clear statement of aims at the top of the webpage. This could include a sentence, or a list of what will be covered on the webpage.
- Thirdly is the inclusion of links to additional information from a diverse range of sources for patients to explore.
- Developers should also be including more information on transparency, references, and currency of information on webpages, something that was inconsistent in the websites reviewed. These should be easy to find on the webpage.

### Limitations

One limitation of this study was the strategy in which finding information regarding Osteogenesis Imperfecta was completed in ChatGPT3.5. It is more likely that patients would search “what is osteogenesis imperfecta” as compared to asking for websites. Aside from the strategy of data collection, ChatGPT3.5 also had limitations of its own. Because the 3.5 version is not updated past September 2021, many of the links provided may have changed after that date, causing 404 errors and hallucinations during data collection (13).

Additionally, the DISCERN assessments were not blinded, both reviewers knew which websites came from either Google, BING, or ChatGPT3.5. This creates potential room for biases in reviewers’ opinions regarding the quality of websites from each search platform. In the future, it would be better to blind this portion of the data collection to reduce any potential intrinsic biases.

## Conclusion

Understanding how health literacy plays into patient care as well as the current websites available to the public through various online platforms such as Google, Bing, and ChatGPT3.5 can help physicians empower their patients and families to make the best educated decisions regarding healthcare and treatment. The observations made in this study could be used in these discussions. Future studies should consider researching the quality of information provided by ChatGPT3.5 that would mimic a true patient experience. Overall, this study was conducted to showcase current gaps in online website education on OI and inform physicians of the quality of websites provided through various online platforms.

## Data Availability

The data is held within a google drive and can be accessed upon request.

## References

1. Rossi V, Lee B, Marom R. Osteogenesis imperfecta - advancements in genetics and treatment. Curr Opin Pediatr. 2019 Dec;31(6):708–15.

2. Deb T. Health Literacy Statistics 2024 By Decisions, Resources… [Internet]. Market.us Media. 2024 [cited 2024 Jul 9]. Available from: https://media.market.us/health-literacy-statistics/

3. DISCERN - The DISCERN Instrument [Internet]. [cited 2024 Jul 9]. Available from: http://www.discern.org.uk/discern_instrument.php

4. Cassidy JT, Baker JF. Orthopaedic Patient Information on the World Wide Web: An Essential Review. The Journal of Bone and Joint Surgery. 2016 Feb 17;98(4):325–38.

5. WebFX [Internet]. [cited 2024 Jul 9]. Readability Test. Available from: https://www.webfx.com/tools/read-able/

6. Johnson D, Goodman R, Patrinely J, Stone C, Zimmerman E, Donald R, et al. Assessing the Accuracy and Reliability of AI-Generated Medical Responses: An Evaluation of the Chat-GPT Model. Res Sq. 2023 Feb 28;rs.3.rs-2566942.

7. Shen Y, Heacock L, Elias J, Hentel KD, Reig B, Shih G, et al. ChatGPT and Other Large Language Models Are Double-edged Swords. Radiology. 2023 Apr;307(2):e230163.

8. Wang G, Gao K, Liu Q, Wu Y, Zhang K, Zhou W, et al. Potential and Limitations of ChatGPT 3.5 and 4.0 as a Source of COVID-19 Information: Comprehensive Comparative Analysis of Generative and Authoritative Information. J Med Internet Res. 2023 Dec 14;25:e49771.

9. Igniting Business [Internet]. [cited 2024 Jul 9]. What is the Proximity Bias (Factor) in Google Search and How Does It Impact SEO (Search Engine Optimization). Available from: https://www.ignitingbusiness.com/blog/what-is-the-proximity-bias-factor-in-google-search-and-how-does-it-impact-seo-search-engine-optimization

10. Alkaissi H, McFarlane SI. Artificial Hallucinations in ChatGPT: Implications in Scientific Writing. Cureus. 15(2):e35179.

11. Osteogenesis Imperfecta (OI) | Boston Children’s Hospital [Internet]. [cited 2024 Jul 9]. Available from: https://www.childrenshospital.org/conditions/osteogenesis-imperfecta

12. Deguchi M, Tsuji S, Katsura D, Kasahara K, Kimura F, Murakami T. Current Overview of Osteogenesis Imperfecta. Medicina (Kaunas). 2021 May 10;57(5):464.

13. GPT-3.5 Turbo Updates | OpenAI Help Center [Internet]. [cited 2024 Aug 23]. Available from: https://help.openai.com/en/articles/8555514-gpt-3-5-turbo-updates

